# ‘People need to be a bit more understanding that my body is wrecked’: A qualitative exploration of inpatient hospital care for people living with multiple long-term conditions

**DOI:** 10.64898/2026.02.08.26345540

**Authors:** Sue Bellass, Thomas Scharf, Miles D Witham, Lynsey Threlfall, Chris Plummer, Avan A Sayer, Rachel Cooper, the ADMISSION Research Collaborative

## Abstract

**Background:** Living with multiple long-term conditions (MLTC) is becoming increasingly common with far-reaching consequences for individuals and healthcare systems. People with MLTC often face complex care pathways through health systems - especially hospitals, which are largely configured for specialist treatment of single conditions - yet evidence on people with MLTC’s lived experience in the hospital setting is limited. This study aimed to understand the hospital care experiences of people living with MLTC who had recently had an inpatient stay.

**Methods:** People with MLTC who had experienced an inpatient stay in hospital within the previous six months were recruited via three hospitals in England and via patient networks. Semi-structured one-to-one interviews were conducted with each participant, focussing on their experiences of care from admission to discharge. An inductive thematic analysis was undertaken.

**Results:** A total of 44 people (mean age 68.4 years, 23 women) who reported living with between 2 and 11 long-term conditions, the majority of whom (96%) reported that their most recent hospital stay was unplanned, participated in the study. Three themes were constructed from the interview data, reflecting perceptions at individual, interpersonal and organisational levels. Participants’ experiences were shaped by *internalised narratives of hospital care*, where care was expected to be focussed primarily on single conditions within a resource-constrained environment. Relationally, the degree of *alignment between clinician and patient knowledge* on conditions was a key contributor to whether hospital care was experienced positively or negatively, and participants’ perceptions of *organisational constraints to holistic care* gave insights into their views on system-level barriers shaping the provision of care for MLTC in the hospital setting.

**Conclusion:** Experiences of inpatient hospital care for people with MLTC are complex, diverse and shaped by expectations of care in a specialist setting configured to provide care for single conditions. Healthcare professionals should incorporate patients’ experiential expertise into decision-making processes through consultation with people with lived experience of MLTC. Redesigning hospital services to provide holistic care will require flexibility to respond to the wide spectrum of MLTC experiences.

## Introduction

Multiple long-term conditions (MLTC), defined as the coexistence of two or more long-term health conditions [1], have far-reaching consequences for both individuals and healthcare systems [2, 3]. Latest estimates suggest that the prevalence of MLTC is high [4], especially in inpatients [5]; for example, 61.0% of adults admitted to hospital in North East England were found to be living with MLTC in 2021-2022, representing a rise since the COVID pandemic which is projected to continue [6–8].

Living with MLTC has myriad adverse effects for individuals and their families related to high symptom and treatment burdens [9], leading to poorer physical function [10] and other adverse outcomes. In addition, evidence that people living with MLTC face challenges navigating fragmented health and social care services is accumulating [11]. People living with MLTC are disadvantaged by healthcare systems configured to provide specialist care for single-organ conditions such as inpatient hospital settings [12], exemplified by their higher rates of inpatient mortality [13] and readmission to hospital [14], longer stays in hospital [15], and lower levels of satisfaction with care [16].

Developing a clearer understanding of people living with MLTC’s experiences of inpatient hospital care is required to inform the transformation of hospital services. Without this it will be difficult to effectively meet the needs of the growing proportion of the population living with MLTC. Recent scoping reviews have summarised the existing knowledge base on perceptions of MLTC hospital care, highlighting the importance of person-centred care and service integration [17, 18]. However, whilst studies have highlighted experiences of hospital care, limited attention has been paid to how experiences are shaped by people’s expectations of care. Further, much of the evidence is from research on comorbid conditions – where another condition (or conditions) occurs alongside and often compounds the effects of an index condition – rather than MLTC, in which no single diagnosis takes primacy [19]. In the UK, qualitative studies of perceptions of hospital care in the context of MLTC have been identified [20, 21], but these studies focus on specific sub-groups (people in the last year of life and pregnant women).

While offering important insights, the existing evidence base therefore has limitations. The heterogeneity of MLTC, and the high prevalence of MLTC among people admitted to hospital, require an understanding of the challenges that MLTC present that go beyond specific comorbidities, for which care pathways may already be established. In this study, nested within a wider research collaborative investigating hospital care and MLTC[22], we aimed to understand how a diverse sample of people living with MLTC experience inpatient hospital care within the National Health Service (NHS) in England, to explore aspects of care that are appreciated by people with MLTC, and to identify priorities for service redesign.

## Methods

A qualitative semi-structured interview study was conducted with people living with MLTC who had experienced at least one inpatient stay in an NHS hospital within the preceding six months. NHS Health Research Authority ethical approval for the study was granted by Wales 3 Research Ethics Committee [reference no. 23/WA/0045].

### Study preparation

The ADMISSION Research Collaborative benefitted from regular advice and guidance from a diverse panel of people living with MLTC and carers (ADMISSION Patient Advisory Group; PAG) from pre-award to the end of the funded period. At the pre-award stage, the PAG highlighted the need for a qualitative study to capture the lived experience of MLTC hospital care [22]. Post-award, discussions and an online survey with the PAG informed the topic guide and study design. In addition, members of the PAG reviewed public-facing documentation and took part in pilot interviews to support refinement of the interview topic guide.

### Sampling and recruitment

Given the highly heterogeneous nature of MLTC, purposive sampling was undertaken to construct a diverse sample of people based on a range of characteristics including age, gender, ethnicity and condition combination. Identification of potential participants was undertaken by hospital clinicians at three NHS sites in England, two in the North East (The Newcastle upon Tyne Hospitals NHS Foundation Trust; Gateshead Health NHS Foundation Trust) and one in the North West (Northern Care Alliance NHS Foundation Trust). Clinicians supporting study recruitment approached potentially eligible patients nearing discharge to request consent for contact by the research team, providing the potential participant with a study information sheet. Recruitment was supplemented by posters in waiting areas at NHS sites, promoting the study via VOICE, a network of public contributors hosted by Newcastle University, and through advertisement in Healthwatch Leeds. The study was also promoted on X (then Twitter) and through personal networks. Several voluntary sector organisations serving groups of people likely to be living with MLTC in the North East and North West of England were also contacted with a request to promote the study across their networks.

Inclusion criteria for the study were: people aged 18 years or older with capacity to consent who were living with any combination of two or more long-term conditions (defined as mental, physical or long-term infectious conditions that had lasted, or were expected to last, for more than a year), and who had experienced at least one inpatient stay of at least one night in an NHS hospital within the last six months. Data collection took place between November 2023 and December 2024. The sample was closely monitored by the research team throughout the recruitment process to identify and address under-represented characteristics. Regular meetings were held with clinicians at recruiting NHS hospital sites to provide updates on the sample and advise of target characteristics.

### Arranging and conducting interviews

Potential participants who had consented to contact by the research team were approached according to their preferred time and method of contact (either telephone or email), which they had been asked to provide on the consent to contact form. The researcher (SB), an experienced post-doctoral qualitative researcher, made three attempts to contact potential participants.

If the potential participant was eligible and willing to take part in the study, the researcher arranged a mutually convenient time for interview, either in-person, via telephone or videoconferencing software to enable the participant to choose their preferred means of engagement. Prior to the interview taking place, consent was given by the participant either via a signed paper consent form or audio recording. An interview topic guide on hospital care, shaped by extant literature and developed through consultation with the research team and the ADMISSION Patient Advisory Group (PAG), was employed flexibly (see Supplementary File 1). Sociodemographic and self-reported health condition data were collected during the interview. No incentives to participate were provided.

All interviews were audio-recorded and transcribed in intelligent verbatim style by a reputable transcription company with extensive experience of transcribing research interviews. Transcripts were checked and anonymised before being imported into NVivo 14 for coding.

### Analysis

Data were analysed inductively using thematic analysis [23, 24]. Following familiarisation with the data, achieved through thorough reading of the transcripts, the researcher generated preliminary codes from five transcripts to develop an initial coding frame, which was then iteratively refined through applying the coding structure to a further five transcripts. This structure was then applied to the data, revising where necessary to capture additional patterns and meanings. Themes and sub-themes were developed through iterative processes which were sense-checked with other authors through discussion at team meetings. The theme structure was then further refined to ensure representation of the dataset.

## Results

Of a total of 92 consent to contact forms or self-referrals received by the research team, 44 participants were consented to the study (Figure 1). Semi-structured interviews were conducted with this diverse sample of 44 adults (23 women) aged 34-87 years old (mean 68.4) who had all experienced an inpatient stay in an NHS hospital within the preceding six months. Twenty-three participants lived in the North East of England, 20 in the North West and one in Yorkshire. Most participants (38/44, 86%) were recruited from NHS sites, with an additional four from VOICE, one via an advertisement in Healthwatch Leeds and one from personal networks.

**Figure 1:**
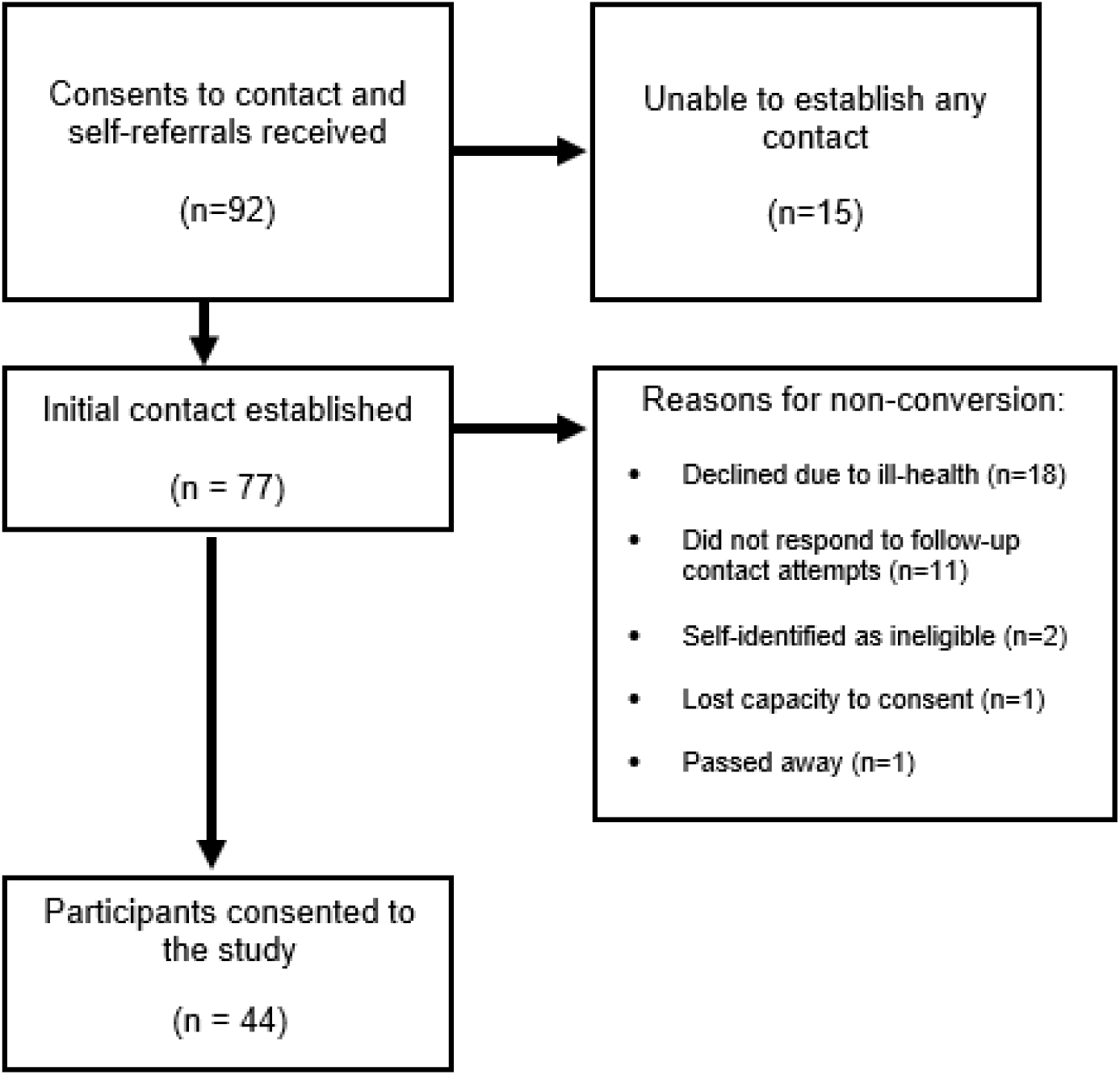
**Participant conversion flow diagram**

Most interviews (29/44, 66%) were conducted in-person in participants’ homes, with 13 (30%) taking place via telephone, and two (4.5%) via videoconferencing software. Interviews lasted between 21 and 129 minutes, with a mean duration of 63.2 minutes.

Participants reported living with between two and 11 long-term conditions. With the support of a clinician, the conditions were grouped according to affected body systems (Supplementary file 2) using a categorisation system developed by members of the research team (MDW, AAS, RC) based on ICD-10 codes [25]. Most participants (42/44, 96%) had experienced an unplanned admission during their most recent hospital stay and had stayed in hospital for less than a week (38/44, 86%) (Table 1). The sample was diverse according to a range of different sociodemographic characteristics (Table 1 and Supplementary file 3).

**Table 1:** Characteristics of study participants.

| Characteristic | Frequency<br>N (%) | Characteristic | Frequency<br>N (%) |
| --- | --- | --- | --- |
| **Gender** |  | **Body system/s (based on ICD-10 chapters) for long-term conditions reported** |  |
| Female | 23 (52) | Cancer | 11 (25) |
| Male | 21 (48) | Cardiovascular disease | 19 (43) |
|  |  | Digestive disease | 8 (18) |
| **Age (y)** |  | Ear disease | 5 (11) |
| 30-39 | 3 (7) | Eye disease | 3 (7) |
| 40-49 | 2 (5) | Haematological disorder | 2 (5) |
| 50-59 | 4 (9) | Infectious disease | 1 (2) |
| 60-69 | 8 (18) | Mental and behavioural disorder | 3 (7) |
| 70-79 | 18 (41) | Metabolic and endocrine disorder | 13 (30) |
| 80-89 | 9 (21) | Musculoskeletal disease | 19 (43) |
| **Index of Multiple deprivation** |  | Neurological disease | 6 (14) |
| Most deprived | 5 (11) | Respiratory disease | 17 (39) |
| 2<sup>nd</sup> | 11 (25) | Urogenital disorder | 7 (16) |
| 3<sup>rd</sup> | 5 (11) |  |  |
| 4<sup>th</sup> | 5 (11) |  |  |
| 5<sup>th</sup> | 1 (2) | **Type of hospital stay** |  |
| 6<sup>th</sup> | 3 (7) | Unplanned | 42 (96) |
| 7<sup>th</sup> | 3 (7) | Planned | 2 (5) |
| 8<sup>th</sup> | 2 (5) |  |  |
| 9<sup>th</sup> | 4 (9) |  |  |
| Least deprived | 4 (9) | **Length of most recent hospital stay** |  |
| No code | 1 (2) | Overnight | 6 (14) |
|  |  | 2-4 nights | 26 (59) |
|  |  | 5-6 nights | 6 (14) |
| **Ethnicity** |  | 1-4 weeks | 3 (7) |
| White British | 43 (98) | More than 4 weeks | 3 (7) |
| White African | 1 (2) |  |  |
| **No. of self-reported long-term conditions** |  | **Inpatient stays in last year** |  |
| 2-3 | 14 (32) | 1 | 22 (50) |
| 4-5 | 18 (41) | 2-4 | 17 (39) |
| 6-7 | 11 (25) | ≥5 | 5 (11) |
| ≥8 | 1 (2) |  |  |

Three interrelated yet distinct overarching themes with sub-themes were developed (Table 2), which reflected perceptions at individual, interpersonal and organisational levels: internalised narratives of NHS hospital care, perceptions of alignment between clinician and patient knowledge, and organisational constraints to holistic care.

**Table 2:** Theme Structure.

| **Themes** | **Sub-themes** |
| --- | --- |
| **Internalised narratives of hospital care** | Hospital as inherently specialised |
|  | Hospital as a resource-constrained environment |
|  | Transparent routines, opaque processes |
| **Alignment between clinician and patient knowledge** | Involvement in the care process |
|  | Medication management |
| **Organisational constraints to holistic care** | Integration of services |
|  | Information continuity |
|  | Perceptions of clinical training and guidelines |

### 1. Internalised narratives of NHS hospital care

Evident throughout the accounts of study participants were the ways in which public narratives of hospital care shaped expectations of the care they would receive. This theme comprised three sub-themes including hospital as inherently specialised, hospital as a resource-constrained environment, and transparent routines but opaque processes.

#### a) Hospital as inherently specialised

Study participants generally understood hospitals to be specialist, short-stay environments, where acute problems would be resolved and they could return to their prior life on discharge. Apart from expecting to receive medication for their long-term conditions, or an acknowledgement of limitations caused by other conditions, participants generally did not anticipate that care would be provided for conditions that had not precipitated their hospital admission. As one participant explained:

“They’ll monitor you if your heart is not working properly because they’ve got the machines, you know what I mean, but you’re specifically just for whatever you go in for. You can’t go in and say, “Can you check my back and my arm?” (GHM01; man, age range 66-70, eleven LTC) Other participants expressed similar views, with one man noting that he had not expected to receive care for another of his health conditions: “It’s a totally separate issue. In other words, we specialise in this, we specialise in that, and that’s all we do” (NCM07, man, age range 81-85, three LTC). Mirroring the single-condition focus of hospitals, participants frequently conceptualised their health biomedically, adopting language relating to body systems, diagnostic labels or specialisms when referring to their inpatient stays, rather than symptoms resulting from LTC.

#### b) Hospital as a resource-constrained environment

Prominent in the accounts were descriptions of hospital as a resource-constrained environment due to a combination of insufficient staffing for the high demand for care. Media reports or personal experience of long waits on hospital corridors in emergency departments were commonly referred to, with participants perceiving beds to be the currency of the hospital care system. This could result in participants questioning the legitimacy and deservingness of their bed occupation based on an assessment of their symptoms. One participant noted, for instance:

“I felt like a fraud, because I didn’t actually feel ill. I just felt very tired and I had a very dry mouth. Those are the only symptoms I had, was my problem. I kept thinking, “Oh, there are all these people coming in that need the bed and I’m sat here just…” (NCF02, woman, age range 71-75, four LTC)

Although this experience is unlikely to be limited to people living with MLTC, having more than one condition could be a complicating factor in the duration of bed occupancy, and the desire to relinquish it. Discharge processes could be delayed, for instance, due to waiting for advice or information from another specialism, leading to participants feeling frustrated by being unable to surrender their bed for someone more unwell. One participant, for instance, noted:

“They said, ‘We’re still waiting for renal,’ and so I just waited and then one of the junior doctors came and I said… I wasn’t rude, I wasn’t rude or anything like but I just said to her, ‘I’m just bed-blocking.’ I said, ‘If renal don’t come down in the next couple of hours, then I’m sorry but I’m going home.’” (NCF01, woman, age range 81-85, six LTC)

The awareness of resource limitations in the hospital setting could impact not only perceptions of legitimate occupation of a bed but also receipt of nursing care. Participants frequently commented on the busyness of staff, particularly nursing staff, who were often sympathetically described as being “rushed off their feet” (NCF06, woman, age range 71-75, six LTC). Some participants felt that this affected the ability to provide person-centred care, with one man noting ‘they’re always too busy for an individual person. “I think, ‘Oh, they don’t care about me’” (NCM03, man, age range 76-80, four LTC), but generally an awareness of the pressures affecting the hospital system shaped the level of care they expected to receive. For example, one man noted:

“Because, you know, if you’ve got another 200 people to look after, they’re not going to get round to you as often as if there were 50 people… they’re not going to come round and, you know, mop my fevered brow or anything, are they? They’ve got a job to do” (NNM02, man, age range 71-75, five LTC)

The sense of busyness could result in a perception that staff did not have capacity to respond to health conditions other than the reason they had been admitted to hospital. Some participants perceived that, due to workload pressure, staff would only respond to conditions if they became unstable during a hospital stay. One participant stated:

“They don’t have time to tie everything together when you’ve got multiple illnesses. It’s just kind of like, “Well we’ll do this and, if anything happens with your other problems then, we’ll deal with that.” (NCM10, man, age range 46-50, four LTC)

#### c) Transparent routines, opaque processes

The third element of this theme contrasts participants’ expectations and experiences of hospital routines with their pathways through the hospital system. Participants frequently described routinised aspects of their time in hospital – monitoring of temperature and blood pressure, the arrival of food and medication, visiting times and night-times – as anticipated and predictable events. However, such routines contrasted with participants not knowing or expecting to know how their journey through the hospital would unfold from admission to discharge. Participants typically expected long waits on hospital corridors, occasionally referring to “horror stories” (NCF09, woman, age range 61-65, three LTC) of such occurrences in the media. Whether they would be admitted to a ward, or remain in an assessment unit, was largely unknown. Although several participants described being asked if they would like to delay discharge for an additional day, in general participants neither knew nor expected to know the timing or stages of their pathway through the hospital system, instead often expressing surprise when processes involving receiving care from more than one department ran smoothly or more quickly than expected:

> “I was in hospital, the following day, having this little op so the transfer from that one department to this went as smooth as anything…That was all within two days…That was a big shock to my system.” (NCF07, woman, age range 86-90, five LTC)

### 2. Alignment between clinician and patient knowledge

Study participants gave both positive and negative accounts of their hospital care, with a key difference between the accounts being whether patients perceived their and the clinicians’ knowledge of their conditions to align. Participants giving a positive narrative generally felt involved in the care process, confident that their medications were well-managed, trusted their clinicians and were able to use their agency to shape decisions about their care. In contrast, people giving negative accounts felt excluded from care decisions, reported being unable to influence the care process, felt their medications were not managed well and expressed doubts in clinicians’ knowledge and skills.

#### a) Involvement in the care process

Key to feeling involved in the care process was shared decision-making during the hospital stay. Participants who described discussing and agreeing treatment options with medical staff held positive views of their care. One man, who has had long-term relationships with clinicians through developing a kidney disorder as a child, noted:

> “Every doctor I speak to, we have a two-way conversation. They listen to what I’m going through. They give me their feedback. And that’s brilliant, and it has worked well.” (NCM04, man, age range 36-40, three LTC)

In contrast, participants reported frustration if they felt that the clinicians were not taking into consideration their other LTC. One participant, for instance, noted that “all the time I was in there I was concerned about my tummy, but nobody else was” (NNF03, woman, age range 76-80, four LTC). Another stated “you’re constantly having to explain that you’ve got sleep apnoea, that you’ve got breathing issues, that you’ve got Crohn’s, that you’ve got problems with your kidneys. Then the next person comes in and you have to go through it all [again].” (NCM10, man, age range 46-50, four LTC).

There were several instances of participants providing information relating to other conditions that they felt were not taken into account by hospital clinicians. One woman, in age range 36-40, attending for an operation, described a situation in which her concerns about incontinence were not responded to, causing frustration and embarrassment:

> “I told people at my pre-assessment…’I have these three conditions. I have Crohn’s disease, I have-‘ I remember telling, at my pre-assessment, that, ‘I struggle with bowel incontinence. So, obviously, most of the time, it’s controlled, but I can’t tell you what all the medication you use in putting me under for surgery is going to do to my poor bowels.’ So, I said, ‘Do you need me to - do you provide incontinence pads at hospital? Should I be wearing one? Should I bring my own in?’ And I remember, when I got to hospital, and I woke up from my surgery, that hadn’t been accounted for.” (NNF02, woman, age range 36-40, three LTC)

Similarly, frustrations could occur when there was a sense that staff would treat all patients who had gone through a particular procedure in the same way, overlooking other LTC that the person lived with. For example, one participant who had had a kidney transplant, noted:

> “Some of them [nurses] were just coming in and being like, ‘You need to get in your chair, you need to get up.’ And I think, when you’ve had your stomach cut open and your body is already weak from everything else you’ve been through, it’s not as easy as someone that has never been through anything before… Whereas when I was there, sometimes I just needed to rest. But I just feel like, with my care, people need to be a bit more understanding that my body is wrecked, and sometimes I might not bounce back as quickly as someone else.” (NTF06, woman, age range 36-40, four LTC)

#### b) Medication management

Care for other LTC in the hospital setting was more frequently achieved through medication management rather than through contact with other specialist teams in the hospital setting. Varying experiences with medication management were reported by study participants. For some, medication for all of their conditions was received in a timely way:

> “I just said, ‘This is what I’m on,’ and they said, ‘Oh, that’s fine, so everything’s okay.’ So, the next thing, I woke up in the morning and, ‘Here are your tablets,’ and, ‘Oh, right, thank you.’” (NCF02, woman, age range 71-75, four LTC)

For other participants, receiving their usual medication for LTC was more problematic. Some reported not receiving the right medication, or receiving it at the wrong time of day, leading them to use medication they had taken into hospital, or to ask family members to bring medication from home when visiting. One person noted:

> “I took them [tablets] in with me because I know I take an awful lot of tablets. They said, ‘Well…’ …I said, ‘What are you giving me them for? I’ve got my own.’ ‘Well we’ve done these now.’ I said, ‘Well you’ve not given me enough.’ I said, ‘I take… Baclofen three times a day’, they were only giving me one. I went, ‘No, I have three so I’m going to take my own.’ Things like that, it’s just… They’re not reading the notes properly, are they?” (NCF03, woman, age range 61-65, four LTC)

Other participants reported similar tensions with staff when self-medicating in the hospital setting. One participant, for instance, living with severe COPD and emphysema amongst other conditions, reported that the staff had not read her records, and that her nebulisers had not been provided:

> “Mistakes were made and I didn’t get all my meds. I mean I was supposed to have my nebs [nebulisers] four times a day. I didn’t get any… Well, fortunately, I have my own portable one… They don’t like using your own equipment. They want you to use theirs, but if they don’t bring it to you, you can’t use it.” (NCF04, woman, age range 71-75, six LTC)

### 3. Organisational constraints to holistic care

Several participants described their perceptions of organisational constraints which impacted holistic, person-centred care in the hospital setting. This theme comprises two sub-themes: integration of services, and perceptions of clinical training and guidelines.

#### a) Integration of services

Some participants described receiving integrated care during an inpatient stay, but the effectiveness of this depended on how well the specialties communicated with each other. For instance, one participant described how his kidney transplant depended on integration between renal and neurology specialists:

> “When I was on dialysis I had to have a high dose of steroids, because of the nerves… But we had to put the transplant back and back, because they couldn’t transplant me on such a high level of steroids. So we had to have a balance between the two conditions of getting the nerves to a point that they were under control, and that they wouldn’t relapse, but the steroid not being too high… so I was quite a unique case from that point of view, because renal and neuro had to really integrate with each [other] on a very regular basis.” (NCM04, man, age range 36-40, three LTC)

Such positive experiences of service integration could engender a sense of safety, where the needs of one condition were optimally balanced against another. However, there could be hidden difficulties integrating services in the hospital setting which create concerns for people living with MLTC, particularly regarding self-management. For example, one participant described difficulties self-managing his diabetes, leading to concerns about safe use of medication:

> “You’re always waiting for a specialist to come and see you. You know? Like, somebody will come for my spleen, and then the nurse will say, ‘Oh, the fella for your spine is coming,’ but it’s quite late in the evening, really, sometimes. So the sheer number of people that deal with me, and my broken body, does make it hard, because you do a lot of waiting. And they might say, ‘Don’t have any lunch until you’ve seen…’ Dr, or Mr. …. But my food has a bearing on when I take my insulin, so then it does become important…because I could be putting more insulin in than my body can stand.” (NCM01, man, age range 71-75, five LTC)

#### b) Information systems

Participants reported difficulties with information transmission within the hospital setting including missing information and medication errors which required repeating their problems to multiple clinicians, occasionally receiving conflicting advice. Difficulties with information sharing could be exacerbated by receiving care for different conditions from two localities:

> “So I’m split across the two boroughs… They work on different systems, so there’s no link. You know, there’s no… consistency with being able to see all your information, so they’ve got a bit on one system, they’ve got a bit on another” (NCM08, man, age range 61-65, three LTC)

While some reported that discharge summaries were comprehensive, others noted a range of problems including discovering unexplained medication changes, inaccurate information, or conditions they were unaware they had been diagnosed with, such as hospital-acquired pneumonia or non-alcoholic fatty liver disease. One woman noted that “all it said was about some tablets which I never received off them anyway” (NCF03, woman, age range 61-65, four LTC), while another participant had been given a summary for a procedure she had undergone the year before. Further, a participant reported a potentially serious information error:

> “And when the letter came, and I got the copy for the GP, it had my name, and all my numbers on the top. It had the change of medication on the bottom, but it turned out to have somebody else’s full details in the middle” (NNM01, man, age range 76-80, three LTC)

#### c) Perceptions of clinical training and guidelines

Some participants perceived that clinicians were limited in their ability to deliver MLTC care due to their training and the clinical guidelines they were expected to use, which were described as “tick box” (NNF03, woman, age range 76-80, four LTC) and could constrain clinicians’ ability to respond flexibly to the needs of individuals in their decision-making processes. One participant noted the limitations of this approach, acknowledging that staff autonomy to make care decisions could be constrained by the organisational context:

> “[Other conditions] are taken into account, but there’s a little bit of a flaw that they’re basically told to follow their certain pathway. Then, if A doesn’t work out, go to B. They follow it until it says no… So they’ll try to bend the rules but, at the end of the day, they’ve got masters to speak to as well so they can only do so much. So you’re listened to but there’s a way of doing it and that’s dictated to the staff.” (NCM10, man, age range 46-50, four LTC).

In addition, participants with rare conditions such as Still’s disease and uncommon tumours remarked on delayed diagnoses and a need for clinicians to have greater training and awareness of the conditions to improve care and to avoid potential interactions between medications.

## Discussion

The findings from this study illuminate the experiences of people living with MLTC when they are admitted to hospital, represented in three themes spanning individual, interpersonal and organisational levels: internalised narratives of hospital care, alignment between clinician and patient knowledge, and organisational constraints on holistic care.

In recognition of the current limitations of siloed healthcare systems [26], much of the qualitative research evidence base on hospital care for people with more than one condition has highlighted the need for greater service integration and holistic person-centred care [27, 28]. By identifying some of the challenges experienced by people living with MLTC receiving inpatient care, the findings from this research echo that work, but also extend it by drawing attention to expectations of episodes of inpatient care, and how these are grounded in the context of hospital narratives of hospital care. Media reports or personal experiences of hospitals as an overwhelmed service, with long waits in Accident and Emergency departments and people receiving treatment on corridors [29], moderated expectations of care in the hospital setting, leading to expressions of surprise when efficient care was encountered.

Aside from medication provision, there was limited expectation that conditions other than the one that had precipitated admission would receive treatment. Inpatient hospital care was generally perceived as a discrete episode, an interruption in the daily flow of self-managing MLTC or seeking support from primary care services. Participants’ perceptions of hospital care were shaped by their experience of MLTC outside the hospital environment. In this regard, the biomedical definition of MLTC - living with more than one condition concurrently - may not fully align with lived experience. As Porter et al. [30] note, the definition of MLTC is predicated on two basic assumptions, firstly that people conceptualise health and illness in relation to diagnostic labels and secondly that health conditions are phenomenologically experienced as co-existing. Porter et al. [30] argue that, rather than a lived experience of concurrence, LTC ‘slip in and out of apprehension according to their perception’. In this study, where the circumstances were such that many other conditions could be managed with medication, participants foregrounded the condition precipitating their admission. Hospital settings, generally understood as providing specialist short-term care for single conditions, may contribute to this foregrounding effect, enabling other conditions to become less visible in comparison to the presenting condition.

There were instances where self-management of LTC could be complicated by a tension between the routines of the hospital setting and the aim of participants to continue to enact their established practices [31, 32], whether through medication, resting when needed or use of incontinence products. Accustomed to self-management in their own homes, and with a tacit knowledge accrued through experience [32], people living with MLTC could feel anxious or frustrated when their medication regimens were not sustained in hospital, sometimes taking in their own medication or acquiring it through family members. Healthcare professionals should be aware of self-management practices to ensure that MLTC care is discussed within the hospital setting, incorporates patients’ experiential expertise into decision-making processes [21, 31] and is clinically safe, avoiding drug-drug interactions.

Although other LTC did not challenge the dominance of the condition leading to admission for all participants in this study, there were clearly circumstances of medical complexity when other conditions were or became clinically significant, and where integration between clinical specialties was needed. In these circumstances, cross-specialty communication was perceived as essential to optimise care delivery, and this study therefore resonates with other research that calls for integrated care models able to respond to the complexity of MLTC [33, 34].

The diversity of experience within our sample highlights the importance of avoiding the assumption that there is ‘one size fits all’ for people living with MLTC who are admitted to hospital. Services need to be flexible enough to respond to this heterogeneity, managing medically complex situations efficiently and acknowledging the experiential knowledge, self-management practices and complex prioritisation processes undertaken by people living with MLTC [35, 36]. Understanding the lived experience of MLTC, as beyond counts of conditions and as phenomenologically inconstant, [32, 35, 36] will promote person-centred, individualised care approaches, the need for which has been highlighted in previous studies of hospital care for MLTC [26, 37].

There are striking parallels between the findings of this study and those of a qualitative study of frontline hospital clinicians’ approaches to decision-making for people with MLTC undertaken within the same research programme [22, 38]. Both studies have identified the tension between care for acute and chronic conditions within the predominantly specialist model of hospital care, resulting in uncertain pathways through the hospital system and inflexible care delivery or receipt. The fundamental misalignment between a system configured to provide single-organ care and the complex needs of people with MLTC entering hospital is exemplified by the constraining effects of single-condition clinical guidelines [39] and compounded by under-staffing in the hospital environment. Embedding generalist skills through clinical training and professional development is, therefore, a crucial step in equipping hospital systems to provide care efficiently for the growing number of people with MLTC [12].

## Strengths and limitations

A key strength of this qualitative study is the exploration of experiences of a relatively large sample of people living with MLTC who had recently received inpatient hospital care. Further, having been collected in 2023 and 2024, the data are contemporaneous and relevant to hospital care in the post-COVID era. The patients in the sample are diverse in several respects, including age, gender, LTC and level of neighbourhood deprivation. Despite their heterogeneity, patterns of experience could be discerned which contribute to understanding the experience of MLTC in English hospitals. Secondly, virtually all of the patients were admitted for unplanned care. Evidence indicates that people with MLTC have increased risk of emergency hospitalisation [40], with associated high costs [41]. Gaining insights into unplanned care is therefore important to address the challenge of reducing emergency admissions.

We acknowledge some limitations. We have less insight into experiences of people living with MLTC admitted to hospital for elective procedures or who stayed in hospital longer than a week. Secondly, while our sample was diverse in many respects, there was a lack of ethnic diversity due to the demographic profile of the areas served by the NHS recruiting sites. Concerted efforts to diversify the sample through other recruitment routes, including contacting voluntary sector agencies serving minority populations, were made but without success. Future research could address these limitations through engaging recruitment sites serving ethnically diverse populations and encouraging recruitment of people with MLTC experiencing planned admissions.

### Policy implications

This study has important policy implications for care provision. With high global prevalence of MLTC [6], healthcare systems worldwide are facing challenges sustaining services originally designed to treat single conditions. In England, for example, an independent investigation has recently described the NHS as being in ‘serious trouble’ [42] (p.1), identifying the inexorable rise in MLTC as a key factor intensifying the pressures on service delivery. In response, the 10 Year Health Plan for England [43] has set out radical transformations needed to create a sustainable health service capable of providing services to current and future populations, including a shift from hospitals to community settings. With this shift, the aim would be for care to be provided at home wherever possible, in neighbourhood health centres when further support is required, and only in hospital if absolutely necessary. Understanding the needs of people with MLTC currently accessing hospital care is an important step towards achieving this goal [5]. Implementing such radical changes in service delivery will need to be communicated effectively so that people living with MLTC are aware of how and where their care will be provided. Incorporating patient groups in planning the rollout of these transformations will be important to achieve this aim. In addition, our data suggest that reconfigured services need to take account of the self-management practices and complex prioritisation processes of people living with MLTC in order to move closer towards delivery of person-centred care.

## Conclusion

Healthcare systems that focus on the delivery of specialised care such as hospitals need transformation in order to address the increasing and complex challenge that MLTC presents. People living with MLTC who are admitted for inpatient hospital stays are highly diverse, with experience that goes beyond counts of conditions to incorporate fluctuating priorities, frustrations and concerns, and experience-based self-management practices. Redesigning hospital services to provide joined-up, person-centred care will require flexibility to respond to the wide spectrum of MLTC experiences. Involving people with lived experience of MLTC in the redesign of these processes is essential to ensure that hospital services can be reconfigured to adequately meet their needs.

## Declarations

### Ethics approval and consent to participate

The study received approval from the Health Research Authority (HRA) and Health and Care Research Wales (HCRW) on 16^th^ March 2023 following review by Wales 3 Research [ref. no. 23/WA/0045]. Consent to participate was given by all study participants prior to interviews being conducted.

### Consent for publication

Not applicable.

### Availability of data and materials

The data generated and analysed during the current study are not currently available due to ongoing follow-up data collection but will be archived in a university-managed dataset in future.

### Authors’ contributions

This study was conducted as part of a programme of research (ADMISSION) conceived by AAS, TS, MDW, CP and RC. SB, TS, MDW and RC conceived the study and developed the protocol. AAS was the Chief Investigator, TS, MDW and RC were Study Supervisors, and SB was the Study Co-ordinator. Under supervision from RC and TS, SB conducted the data collection and analysis. All authors contributed to the interpretation of data. SB wrote the first draft of the manuscript, supported by RC. All authors critically revised subsequent iterations of the manuscript and approved the final version.

## Acknowledgements

The authors wish to acknowledge the ADMISSION Research Collaborative’s Patient Advisory Group (PAG)— a diverse group of people living with multiple long-term conditions and carers who supported the research team from pre-award through to the end of the funded period. Members of the PAG were involved in this qualitative study in several ways, including contributing to the interview topic guide, reviewing project documentation, advising on aspects of study design, and participating in pilot interviews.

We would like to acknowledge other members of the recruiting teams in the three local NHS sites. Newcastle upon Tyne Hospitals NHS Foundation Trust: Alaa Ebraheem; Gateshead Health NHS Foundation Trust: Claire McDonald, Ian Sayers and Bryony Storey; Northern Care Alliance NHS Foundation Trust: Emma Vardy, Louise Tomkow, Bethan Charles, Tahira Dawoodji, Ashliegh Lovett, Akifah Patwary and Jane Shaw.

We would also like to acknowledge Jonathan Bunn who classified study participants’ self-reported health conditions in alignment with the ADMISSION Research Collaborative’s approach to defining and operationalising MLTC (https://doi.org/10.1007/s41999-024-00953-8)

Finally, we would like to thank all the study participants for sharing their experiences of NHS hospital care.

## References

1. Academy of Medical Sciences, Multiple long-term conditions (multimorbidity): a priority for global health research. 2018, Academy of Medical Sciences: London.

2. Whitty, C.J., Harveian Oration 2017: Triumphs and challenges in a world shaped by medicine. Clin Med (Lond), 2017. 17(6): p. 537–544.

3. Whitty, C.J., Chief medical officer’s annual report 2023: Health in an ageing society, Department of Health and Social Care, Editor. 2023, UK Government: London.

4. Valabhji, J., et al., Prevalence of multiple long-term conditions (multimorbidity) in England: a whole population study of over 60 million people. Journal of the Royal Society of Medicine, 2024. 117(3): p. 104–117.

5. Cooper, R., et al., A National Health Service in Serious Trouble: what do multiple long-term conditions tell us about deterioration in health among people accessing hospital care in the North East of England? . medRxiv, 2025.

6. Chowdhury, S.R., et al., Global and regional prevalence of multimorbidity in the adult population in community settings: a systematic review and meta-analysis. EClinicalMedicine, 2023. 57: p. 101860.

7. Kingston, A., et al., Projections of multi-morbidity in the older population in England to 2035: estimates from the Population Ageing and Care Simulation (PACSim) model. Age and Ageing, 2018. 47(3): p. 374–380.

8. Watt, T.R., et al., Health in 2040: projected patterns of illness in England. 2023, The Health Foundation: London.

9. Holland, E., et al., The impact of living with multiple long-term conditions (multimorbidity) on everyday life – a qualitative evidence synthesis. BMC Public Health, 2024. 24(1): p. 3446.

10. Aubert, C.E., et al., Multimorbidity and long-term disability and physical functioning decline in middle-aged and older Americans: an observational study. BMC Geriatrics, 2022. 22(1): p. 910.

11. O’Callaghan, C.A., et al., Integrating and Defragmenting Multi-Specialty Care for People With Multiple Long-Term Conditions. British Journal of Hospital Medicine, 2025. 86(8).

12. Whitty, C.J.M., et al., Rising to the challenge of multimorbidity. BMJ, 2020. 368: p. l6964.

13. Almagro, P., et al., Multimorbidity gender patterns in hospitalized elderly patients. Plos One, 2020.

14. Rodrigues, L.P., et al., Association between multimorbidity and hospitalization in older adults: systematic review and meta-analysis. Age and Ageing, 2022. 51(7).

15. Aubert, C.E., et al., Association of patterns of multimorbidity with length of stay: A multinational observational study. Medicine, 2020. 99(34).

16. Hewitson, P., et al., People with limiting long-term conditions report poorer experiences and more problems with hospital care. BMC Health Services Research, 2014. 14(1): p. 33.

17. Bellass, S., et al., Experiences of hospital care for people with multiple long-term conditions: a scoping review of qualitative research. BMC Med, 2024. 22(1): p. 25.

18. Thompson, F., et al., Perceptions of Hospital Care Quality According to People Living With Multiple Long-Term Conditions: A Scoping Review. Health Expect, 2025. 28(3): p. e70297.

19. Khunti, K., H. Sathanapally, and P. Mountain, Multiple long term conditions, multimorbidity, and co-morbidities: we should reconsider the terminology we use. BMJ, 2023. 383: p. p2327.

20. Mason, B., et al., ’My body’s falling apart.’ Understanding the experiences of patients with advanced multimorbidity to improve care: serial interviews with patients and carers. BMJ Support Palliat Care, 2016. 6(1): p. 60–5.

21. Hanley, S.J., et al., Lost in the System: Responsibilisation and Burden for Women With Multiple Long-Term Health Conditions During Pregnancy. Health Expectations, 2024. 27(3): p. e14104.

22. Witham, M.D., et al., Building ADMISSION - A research collaborative to transform understanding of multiple long-term conditions for people admitted to hospital. J Multimorb Comorb, 2025. 15: p. 26335565251317940.

23. Braun, V. and V. Clarke, Using thematic analysis in psychology. Qualitative Research in Psychology, 2006. 3(2): p. 77–101.

24. Green, J. and N. Thorogood, Qualitative Methods for Health Research. 2nd ed. 2004, London: Sage Publications.

25. Cooper, R., et al., Rising to the challenge of defining and operationalising multimorbidity in a UK hospital setting: the ADMISSION research collaborative. European Geriatric Medicine, 2024. 15(3): p. 853–860.

26. Schiøtz, M.L., et al., Quality of care for people with multimorbidity – a case series. BMC Health Services Research, 2017. 17(1): p. 745.

27. Verhoeff, M., et al., Secondary care experiences of patients with multiple chronic conditions. Neth J Med, 2018. 76(9): p. 397–406.

28. Bosire, E.N., Patients’ Experiences of Comorbid HIV/AIDS and Diabetes Care and Management in Soweto, South Africa. Qual Health Res, 2021. 31(2): p. 373–384.

29. Wise, J., Scale of NHS’s “corridor care” is revealed in Royal College of Nursing report. BMJ 2025. 388:r99.

30. Porter, T., B.N. Ong, and T. Sanders, Living with multimorbidity? The lived experience of multiple chronic conditions in later life. Health, 2020. 24(6): p. 701–718.

31. Bratzke, L.C., et al., Self-management priority setting and decision-making in adults with multimorbidity: A narrative review of literature. International Journal of Nursing Studies, 2015. 52(3): p. 744–755.

32. Coventry, P.A., et al., Living with complexity; marshalling resources: a systematic review and qualitative meta-synthesis of lived experience of mental and physical multimorbidity. BMC Family Practice, 2015. 16(1): p. 171.

33. Struckmann, V., et al., Relevant models and elements of integrated care for multi-morbidity: Results of a scoping review. Health Policy, 2018. 122(1): p. 23–35.

34. Looman, W., et al., Drivers of successful implementation of integrated care for multi-morbidity: Mechanisms identified in 17 case studies from 8 European countries. Social Science & Medicine, 2021. 277: p. 113728.

35. Bayliss, E.A., et al., Processes of care desired by elderly patients with multimorbidities. Family Practice, 2008. 25(4): p. 287–293.

36. Lindsay, S., Prioritizing illness: lessons in self-managing multiple chronic diseases. Canadian Journal of Sociology, 2009. 34(4): p. 983–1002.

37. Kuluski, K., et al., The care delivery experience of hospitalized patients with complex chronic disease. Health Expect, 2013. 16(4): p. e111–23.

38. Pretorius, S., et al., Front-Line Decision-Making: A Thematic Analysis of Interviews with Hospital Staff on Referrals, Admissions, and Care for People with Multiple Long-Term Conditions., 2026. **In preparation**.

39. Pretorius, S., et al., Making clinical guidelines work for people living with multiple long-term conditions: Analysis and recommendations from a review of single-condition guidelines. BMJ Medicine, 2026. In press.

40. Dodds, R.M., et al., Simple approaches to characterising multiple long-term conditions (multimorbidity) and rates of emergency hospital admission: Findings from 495,465 UK Biobank participants. Journal of Internal Medicine, 2023. 293(1): p. 100–109.

41. Stokes, J., et al., Multimorbidity combinations, costs of hospital care and potentially preventable emergency admissions in England: A cohort study. PLoS Med, 2021. 18(1): p. e1003514.

42. Darzi, A., Independent Investigation of the National Health Service in England., Open Government Licence, Editor. 2025: London.

43. Department of Health and Social Care, Fit for the future: 10 Year Health Plan for England. 2025, Department of Health and Social Care: London.

