## Supplementary file 1 for "‘People need to be a bit more understanding that my body is wrecked’: A qualitative exploration of inpatient hospital care for people living with multiple long-term conditions"

**Supplementary file 1: Interview Topic Guide**

**HOSPITAL CARE**

- Experiences of admission, stay and discharge: accessing care, perceptions of why they were admitted, length of stay, care pathways/ transitions, hospital routines, experiences of discharge and follow up, perceptions of good and poor quality care.
- Perceptions of the physical space/ hospital environment
- Perceptions of interface between different specialties in secondary care e.g. information sharing and multidisciplinary working
- Perceptions of cross-sectoral care management, co-ordination and continuity: interface between primary and secondary care, and between health and social care systems (and possibly other services such as the third sector/ community organisations)
- Perceptions of knowledge of multiple long-term conditions (MLTC) among health (and social) care professionals
- Perceptions of advice, education or guidance provided by care professionals
- Relationships with care professionals
- Suggestions for ways to improve care

**LIVING WITH MLTC**

- A typical day / week
- Onset of MLTC in the life course and other major life events
- Growing older with MLTC
- Effects of MLTC on sense of self, routines, relationships, neighbourhood/ community life, work, travel, hobbies, social and leisure activities
- Support from others
- Caring responsibilities for others

**DEMOGRAPHIC INFORMATION**

- Age
- Gender
- Ethnicity
- Level of educational attainment
- Current/ previous employment
- Marital status
- Household size/ composition
- Self-report of health conditions /disability
