## Supplementary file 2 for "‘People need to be a bit more understanding that my body is wrecked’: A qualitative exploration of inpatient hospital care for people living with multiple long-term conditions"

**Supplementary file 2 : Number of self-reported health conditions, number of hospital stays in the last year and body systems affected by conditions (according to ICD-10 chapters)**

| **Partici-pant**  **ID code** | **Age range** | **No. of self-reported conditions** | **No. of hospital stays in last year** | **Cancer** | **Cardio-vascular disease** | **Digestive**  **Disease** | **Ear**  **disease** | **Eye**  **disease** | **Haema-tological**  **disorder** | **Infectious**  **disease** | **Mental / behaviour-al disorder** | **Metabolic/**  **endocrine**  **disorder** | **Musculo-skeletal**  **disease** | **Neuro-**  **logical**  **disease** | **Respir-atory disease** | **Urogenital disorder** |
| --- | --- | --- | --- | --- | --- | --- | --- | --- | --- | --- | --- | --- | --- | --- | --- | --- |
| GHM01 | 66-70 | 11 | >=5 |  |  |  |  |  |  |  |  |  |  |  |  |  |
| GHM02 | 71-75 | 3 | >=5 |  |  |  |  |  |  |  |  |  |  |  |  |  |
| NCF01 | 81-85 | 6 | 2-4 |  |  |  |  |  |  |  |  |  |  |  |  |  |
| NCF02 | 71-75 | 4 | 1 |  |  |  |  |  |  |  |  |  |  |  |  |  |
| NCF03 | 61-65 | 4 | 1 |  |  |  |  |  |  |  |  |  |  |  |  |  |
| NCF04 | 71-75 | 6 | 2-4 |  |  |  |  |  |  |  |  |  |  |  |  |  |
| NCF05 | 76-80 | 2 | 1 |  |  |  |  |  |  |  |  |  |  |  |  |  |
| NCF06 | 71-75 | 6 | 1 |  |  |  |  |  |  |  |  |  |  |  |  |  |
| NCF07 | 86-90 | 5 | 1 |  |  |  |  |  |  |  |  |  |  |  |  |  |
| NCF08 | 76-80 | 4 | 1 |  |  |  |  |  |  |  |  |  |  |  |  |  |
| NCF09 | 61-65 | 3 | 1 |  |  |  |  |  |  |  |  |  |  |  |  |  |
| NCF10 | 55-60 | 6 | 1 |  |  |  |  |  |  |  |  |  |  |  |  |  |
| NCM01 | 71-75 | 5 | 2-4 |  |  |  |  |  |  |  |  |  |  |  |  |  |
| NCM02 | 76-80 | 6 | 2-4 |  |  |  |  |  |  |  |  |  |  |  |  |  |
| NCM03 | 76-80 | 4 | 1 |  |  |  |  |  |  |  |  |  |  |  |  |  |
| NCM04 | 36-40 | 3 | 1 |  |  |  |  |  |  |  |  |  |  |  |  |  |
| NCM05 | 81-85 | 4 | 2-4 |  |  |  |  |  |  |  |  |  |  |  |  |  |
| NCM06 | 71-75 | 4 | 2-4 |  |  |  |  |  |  |  |  |  |  |  |  |  |
| NCM07 | 81-85 | 3 | 2-4 |  |  |  |  |  |  |  |  |  |  |  |  |  |
| NCM08 | 61-65 | 3 | 1 |  |  |  |  |  |  |  |  |  |  |  |  |  |
| NCM09 | 71-75 | 3 | 1 |  |  |  |  |  |  |  |  |  |  |  |  |  |
| NCM10 | 46-50 | 4 | 1 |  |  |  |  |  |  |  |  |  |  |  |  |  |
| NNF01 | 61-65 | 2 | 2-4 |  |  |  |  |  |  |  |  |  |  |  |  |  |
| NNF02 | 36-40 | 3 | 2-4 |  |  |  |  |  |  |  |  |  |  |  |  |  |
| NNF03 | 76-80 | 4 | 1 |  |  |  |  |  |  |  |  |  |  |  |  |  |
| NNF04 | 71-75 | 6 | 1 |  |  |  |  |  |  |  |  |  |  |  |  |  |
| NNM01 | 76-80 | 3 | 2-4 |  |  |  |  |  |  |  |  |  |  |  |  |  |
| NNM02 | 71-75 | 5 | 2-4 |  |  |  |  |  |  |  |  |  |  |  |  |  |
| NTF01 | 76-80 | 6 | 1 |  |  |  |  |  |  |  |  |  |  |  |  |  |
| NTF02 | 81-85 | 6 | 2-4 |  |  |  |  |  |  |  |  |  |  |  |  |  |
| NTF03 | 76-80 | 5 | 1 |  |  |  |  |  |  |  |  |  |  |  |  |  |
| NTF04 | 66-70 | 5 | 2-4 |  |  |  |  |  |  |  |  |  |  |  |  |  |
| NTF05 | 31-35 | 3 | 1 |  |  |  |  |  |  |  |  |  |  |  |  |  |
| NTF06 | 36-40 | 4 | >=5 |  |  |  |  |  |  |  |  |  |  |  |  |  |
| NTF07 | 56-60 | 4 | 2-4 |  |  |  |  |  |  |  |  |  |  |  |  |  |
| NTF08 | 46-50 | 2 | 2-4 |  |  |  |  |  |  |  |  |  |  |  |  |  |
| NTF09 | 51-55 | 5 | 1 |  |  |  |  |  |  |  |  |  |  |  |  |  |
| NTM01 | 81-85 | 6 | >=5 |  |  |  |  |  |  |  |  |  |  |  |  |  |
| NTM02 | 71-75 | 6 | 2-4 |  |  |  |  |  |  |  |  |  |  |  |  |  |
| NTM03 | 71-75 | 4 | 2-4 |  |  |  |  |  |  |  |  |  |  |  |  |  |
| NTM04 | 86-90 | 3 | 1 |  |  |  |  |  |  |  |  |  |  |  |  |  |
| NTM05 | 71-75 | 3 | 1 |  |  |  |  |  |  |  |  |  |  |  |  |  |
| NTM06 | 61-65 | 4 | 1 |  |  |  |  |  |  |  |  |  |  |  |  |  |
| NTM07 | 51-55 | 6 | >=5 |  |  |  |  |  |  |  |  |  |  |  |  |  |
