## Supplementary file 3 for "‘People need to be a bit more understanding that my body is wrecked’: A qualitative exploration of inpatient hospital care for people living with multiple long-term conditions"

**Supplementary file 3: Additional demographic characteristics of study participants**

| **Characteristic** | **Frequency**  **N (%)** | **Characteristic** | **Frequency**  **N (%)** |
| --- | --- | --- | --- |
| **Marital status** |  | **Household composition** |  |
| Married or co-habiting | 25 (57) | Lives with spouse/ partner | 18 (45) |
| Widowed | 9 (20) | Lives with spouse and adult child/children | 5 (11) |
| Divorced | 6 (14) | Lives with spouse and young child/ children | 2 (5) |
| Single | 4 (9) | Lives alone | 16 (36) |
|  |  | Lives with adult child/children | 2 (5) |
|  |  | Lives with young child/ children | 1 (2) |
| **Current / previous employment** |  | **Educational attainment** |  |
| Senior professional / manager (e.g. architect, lawyer, director)   1. Retired 2. Employed | 7 (16)  1 (2) | Left school before age 16  GCSEs or equivalent  A levels  Further education qualification | 8 (18)  8 (18)  2 (5)  13 (30) |
| Skilled professional (e.g. nurse, police officer, teacher, electrician)   1. Retired 2. Employed | 10 (23)  2 (5) | Undergraduate degree or equivalent  Postgraduate degree (Master’s) | 11 (25)  2 (5) |
| Manual worker (e.g. driver, coal miner)   1. Retired 2. Employed | 11 (25)  0 (0) |  |  |
| Administrator   1. Retired 2. Employed | 10 (23)  1 (2) |  |  |
| Retired (other) | 1 (2) |  |  |
| Unemployed | 1 (2) |  |  |
